# Wireless Monitoring of Patients Admitted to Hospital Wards: A Stepped-Wedge Cluster Randomized Trial (i-PROTECT Trial): Clinical Study Protocol and Statistical Analysis Plan

**DOI:** 10.64898/2026.09.30.26364373

**Authors:** Yaseen Arabi, Abdulmohsen Alsaawi, Tariq Al Debasi, Ali Al Khathaami, Khalil Almajed, Hasan M. Al-Dorzi, Nabiha Tashkandi, Raed Al Hazme, Amal Matroud, Angela Caswell, Husam I. Ardah, Katja Kjaer Groenbaek, Nouf Al Ayadhi, Abdulaleem Alatassi, Sara Suliman Aldekhyl, Samah Y. Alqahtani, Saad Al-Qahtani, Abdulrahman Al Rashid, Zeyad Yousef, Abdulrahman Alhadab, Roya Alotaibi, Welhemina Molala, Mashail Saud Khadhran Alrashidi, Mawaddah Musallam Alotaibi, Ghada Helal Alenazi, Reem Alotaibi, Bijesh Yadav, Musharaf Sadat, Khalid Alshurman, Abdullah Al Enezi, Wejdan Al Rassamah, Abdulrahman Saleh Al Shuail, Abdulaziz Abdulaal, Abdulaziz Al-Dawood

## Abstract

**Introduction:** Clinical deterioration in patients admitted to hospital wards is a major cause of preventable morbidity and mortality and is frequently preceded by abnormalities in vital signs that occur hours before serious adverse events occur. Early identification of patients at risk provides an opportunity for timely intervention before irreversible organ dysfunction develops. The objective of this trial is to evaluate whether implementing a wireless patient monitoring system reduces mortality and cardiopulmonary failure in patients admitted to hospital wards.

**Methods and analysis:** The i-PROTECT trial is designed as a stepped-wedge cluster randomized controlled trial. Hospital wards (total of 28 wards, constituting clusters in this design) are randomized to have wireless monitoring for selected high-risk patients, 7 wards at a time, with every 7 wards constituting a sequence. In the first period, all wards will have no wireless monitoring. After a baseline period of 2 (+1 transition) months, the intervention is implemented in a randomly selected new sequence every 3-month period until the intervention is implemented in all sequences. The control group will receive usual care with standard intermittent monitoring and no wireless monitoring. The calculated sample size is 13160 patients. The primary endpoint is cardiopulmonary failure; a composite endpoint of 30-day in-hospital mortality, cardiac arrest, and need for vasopressor or intubation. The trial started on Feb 1, 2026, and is expected to complete patient follow-up by the end of May 2027.

**Ethics and dissemination:** The trial was approved by the Institutional review board (IRB) of Ministry of National Guard Health Affairs (IRB/2623/23). The results will be presented at relevant national and international critical care conferences and published in international peer-reviewed journals.

**Clinicaltrial.gov:** NCT06644599

**Strengths and limitations of this study:**

- The pragmatic design reflects routine clinical practice and therefore enhances external validity.
- The study has a large sample size, standardized electronic data collection, and centralized monitoring, minimizing selection bias and missing outcome data.
- The main limitation is the inability to blind healthcare providers and secular changes in clinical practice over time.

## Introduction

Clinical deterioration in patients admitted to hospital wards is a major cause of preventable morbidity and mortality and is frequently preceded by abnormalities in vital signs that occur hours before serious adverse events occur.[1, 2] It encompasses progressive physiological deterioration resulting from acute illness or treatment-related complications and is influenced by multiple factors, including frailty, premorbid functional status, comorbidity burden, severity of illness, response to therapy, and delays in recognition and escalation of care.[1] Early identification of patients at risk provides an opportunity for timely intervention before irreversible organ dysfunction develops and has the potential to reduce major adverse events, including cardiac arrest and death. [3-5] In an analysis of preventable in-hospital deaths, inadequate clinical monitoring, diagnostic errors, and inappropriate fluid or medication management were identified as the most common contributory factors. These findings emphasize that failure to recognize deterioration remains an important patient safety challenge.[5]

Recognition of deteriorating patients is not dependent solely on physiological monitoring but requires an integrated chain of surveillance, interpretation, communication, and timely clinical response.[6, 7] Although many hospitals have implemented rapid response systems and early warning scores such as the National Early Warning Score (NEWS-2), routine intermittent measurement of vital signs every four to eight hours may fail to identify physiological abnormalities that develop between scheduled observations.[8] Consequently, clinically important deterioration may remain undetected until patients have already progressed to advanced respiratory or cardiovascular failure, thereby limiting opportunities for early therapeutic intervention.

Recent advances in wireless technologies have enabled continuous monitoring of physiological parameters on general hospital wards. These systems provide continuous measurement of vital signs regardless of the location and have demonstrated acceptable accuracy, feasibility, and high levels of acceptance among both patients and healthcare providers.[9-11] Continuous monitoring has the potential to identify deterioration earlier than conventional intermittent observations, particularly among patients at increased risk for clinical deterioration.[10] Moreover, integrating continuous physiological monitoring with intelligent signal-processing algorithms and structured clinical escalation pathways may improve alert specificity while reducing alarm fatigue, one of the principal barriers to implementation and early deterioration detection.[12]

Evidence supporting the clinical effectiveness of continuous ward monitoring, however, remains limited. Most published studies have been observational or quasi-experimental and have suggested reductions in mortality, cardiac arrest, rescue events, ICU transfers, or time to treatment following implementation of continuous monitoring systems.[13-17] More recently, a propensity-matched analysis involving nearly 8,000 medical ward patients demonstrated substantially lower rates of ICU transfer or death among patients receiving continuous monitoring compared with standard intermittent monitoring.[17] Although encouraging, these studies remain susceptible to confounding and secular trends inherent to non-randomized designs.

Randomized evidence remains relatively scarce and has primarily focused on postoperative surgical populations. A pilot randomized trial demonstrated earlier antibiotic administration for sepsis, shorter hospital stay, and fewer readmissions among patients receiving continuous remote monitoring.[18] Subsequent randomized trials showed reductions in hypoxemic burden and some adverse clinical events but did not consistently demonstrate improvements in major patient-centered outcomes.[19, 20] Furthermore, most previous trials evaluated single physiological variables, postoperative populations, or simple threshold-based alerting systems rather than integrated monitoring strategies incorporating continuous surveillance, advanced algorithmic analysis, centralized clinical review, and standardized escalation pathways.

Consequently, important uncertainties remain regarding whether continuous wireless monitoring can improve clinically meaningful outcomes in heterogeneous hospital ward populations receiving contemporary standard care.

The Integrated Proactive Wireless Monitoring for Clinical Deterioration (i-PROTECT) Trial was designed to address this important evidence gap. Using a pragmatic stepped-wedge cluster randomized design, the trial evaluates whether implementation of an integrated continuous wireless monitoring pathway for high-risk ward patients reduces cardiopulmonary failure and 30-day in-hospital mortality compared with standard intermittent monitoring. Unlike previous studies that primarily evaluated monitoring devices in isolation, the i-PROTECT intervention combines continuous physiological monitoring, intelligent algorithmic processing, centralized expert surveillance, structured communication, and predefined escalation protocols within an established rapid response system, thereby evaluating the effectiveness of a comprehensive digital care pathway in routine clinical practice. We hypothesize that implementation of such an integrated pathway for high-risk ward patients will reduce the incidence of cardiopulmonary failure and 30-day in-hospital mortality compared with standard intermittent monitoring.

## Methods and analysis

This protocol was developed and reported in accordance with the Standard Protocol Items: Recommendations for Interventional Trials (SPIRIT) 2013 Statement, with the design and reporting of stepped-wedge–specific elements guided by the CONSORT extension for stepped-wedge cluster randomized trials.[21, 22] The intervention is described in accordance with the Template for Intervention Description and Replication (TIDieR) checklist to facilitate transparent reporting and reproducibility of this complex healthcare intervention.[23] Any modifications to workflow, alert thresholds, staffing model, or escalation procedures introduced during implementation will be prospectively documented with justification and timing.

### Settings

The study is conducted at King Abdulaziz Medical City-Riyadh (KAMC-R), Saudi Arabia. The hospital has an integrated electronic medical record (EMR) system, BESTCare, which has been implemented as a joint venture with the Seoul National University Bundang Hospital, South Korea.[24] Its critical applications include clinical documentation, computerized physician order entry, clinical decision support system, and clinical data warehouse.[24] The system is also interfaced with the hospital’s vital signs measurement devices. The WARD algorithm and the Honeywell CT30 mobile alerting infrastructure (see below) are hosted on-premises within the hospital’s data center rather than in an external or vendor-hosted cloud environment, consistent with the institution’s data governance requirements for continuous physiological data. Device-to-EMR connectivity relies on Health Level Seven (HL7) messaging, through which the Masimo Root system transmits observation/result (ORU) messages containing continuous vital sign data directly into BESTCare, while BESTCare in turn provides admission-discharge-transfer (ADT) data to the WARD algorithm to establish patient context. This HL7-based architecture is consistent with recognized approaches for integrating third-party physiological monitoring devices into hospital EMR systems without requiring replacement of existing monitoring infrastructure.[25] [26] KAMC-R has a well-established Rapid response team (RRT) system in place.[3] In addition, all wards have NEWS-2 system [27] and sepsis screening system implemented before the trial. [28]

### Study design

The trial is designed as a stepped-wedge cluster RCT. Hospital wards (which constitute clusters in this design) are randomized to have wireless monitoring, 7 wards at a time, with every 7 wards constituting a sequence of treatment (Figure 1). The study consists of five 2-month periods; a 1-month transition period follows each of the first four periods, during which the next sequence of 7 wards crosses over to the intervention, for a total study duration of 15 months. In the first period, all wards will have no wireless monitoring. After a baseline period of 2 months (+1 month of transition), the intervention is implemented in a randomly selected new sequence of treatment every 3-month period until the intervention is implemented in all wards. This design is suitable for quality improvement projects and incorporates comparisons between the intervention group and the control group, both horizontally (intervention clusters and control clusters at the same time) and vertically over time (the same cluster before and after the intervention). The study was approved by the Ministry of National Guard Health Affairs Institutional Review Board, Riyadh, Saudi Arabia (IRB/2623/23).).

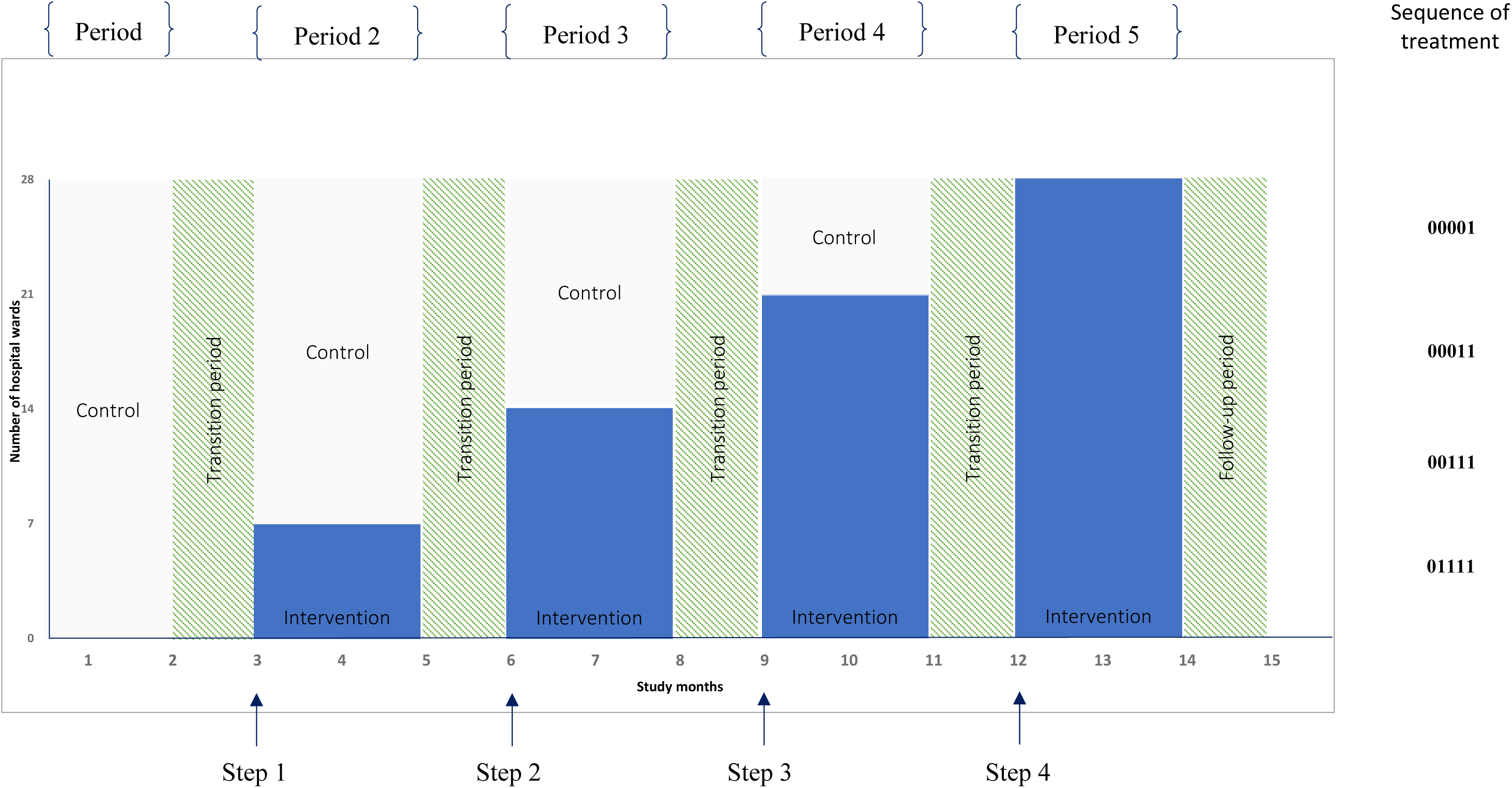

### Inclusion and Exclusion Criteria

#### Ward-level inclusion and exclusion criteria

Inclusion Criteria:

Inpatient wards, defined as wards used to manage adult inpatients.

Exclusion Criteria:

1. Cardiology, pediatric, obstetric wards
2. ICUs and emergency departments
3. Operating rooms
4. Outpatient clinics
5. Daycare wards, endoscopy, outpatient procedure areas, hemodialysis units

#### Patient-level inclusion and exclusion criteria

Inclusion Criteria:

1. Adults as defined by hospital policy of 16 years or older
2. Checked in as inpatient status to one of the study wards

Exclusion Criteria:

Patients with a Do-Not-Resuscitate (DNR) order, or no commitment to full life support, at the time of ward arrival.

### Study Procedures

#### Intervention group

The intervention consists of an integrated clinical workflow that combines continuous physiological monitoring, automated alert generation, centralized clinical surveillance, and a structured escalation pathway (Figure 2). Patients at high risk for clinical deterioration will receive continuous wireless physiological monitoring using the Masimo Root Monitoring System (Masimo Corp., Irvine, CA, USA, Medical National listing No: ME0000002404SFDAA00057). [29] This system continuously measures heart rate, respiratory rate, and oxygen saturation (SpO₂) in addition to intermittently measuring blood pressure. Physiological data are transmitted continuously for centralized monitoring and real-time algorithmic analysis (See below). Patients are eligible for monitoring if they meet any of the following criteria: (1) a NEWS-2 of 5 or higher; (2) a lower NEWS-2 score with clinical concern identified by the treating team; (3) activation of the RRT, regardless of NEWS-2 score; or (4) discharge from the intensive care unit (ICU), regardless of NEWS-2 score.

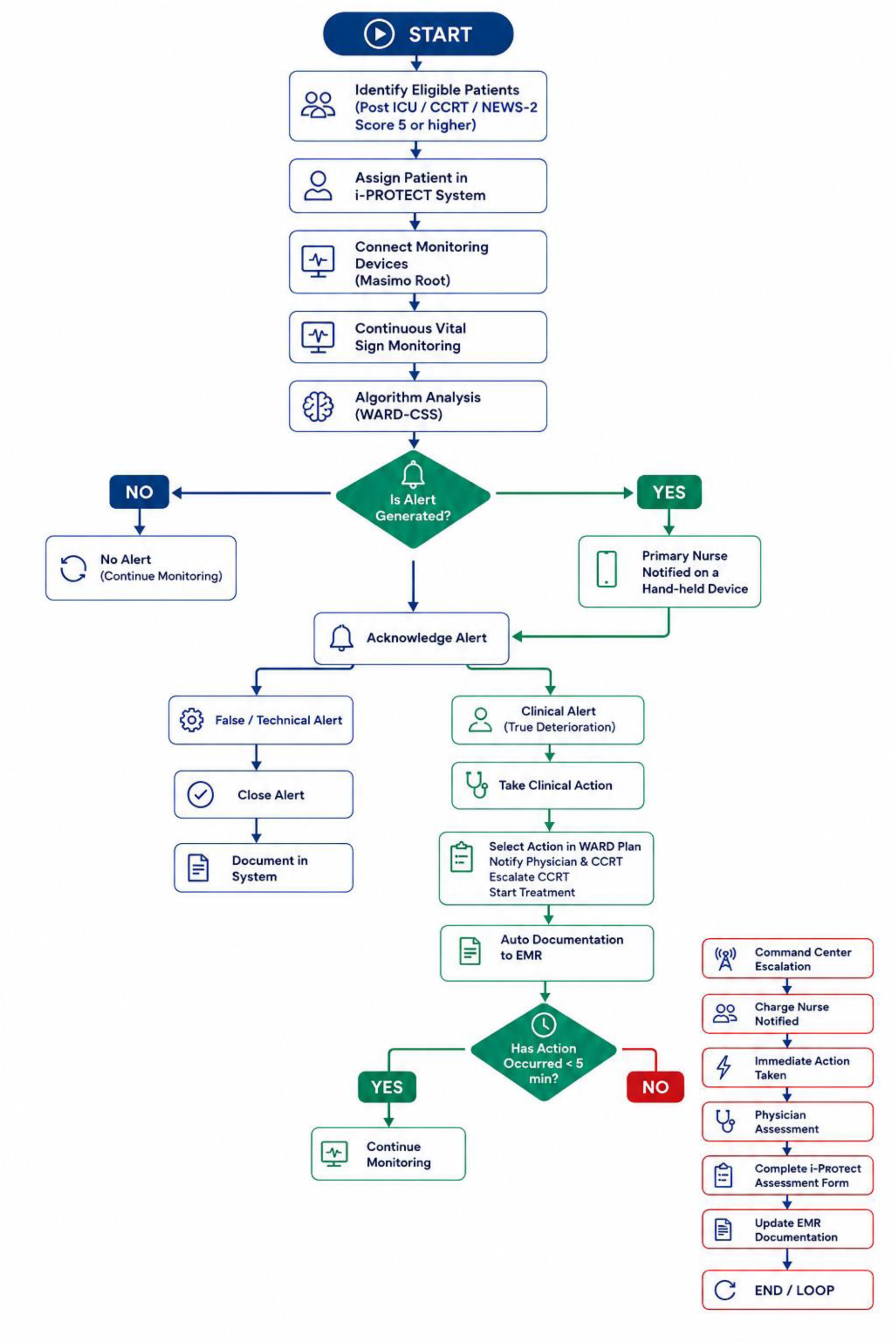

#### Algorithmic processing of physiological data

Continuous physiological data are analyzed using a clinical decision support system (WARD 24/7 ApS, Copenhagen, Denmark; Saudi FDA approval MDMA-2-2026-0643).[20, 30] Rather than generating an alert whenever a physiological threshold is exceeded, the WARD algorithm continuously evaluates vital sign deviations using a proprietary alerting bundle of thresholds and durations before generating an alert designed to maximize clinically meaningful alerts while minimizing false or non-actionable alarms.

The algorithm incorporates several complementary processing steps. First, signal-processing filters reduce noise and measurement artifacts, including transient motion artifacts and temporary sensor disconnections. Second, physiological abnormalities are evaluated according to both their severity and persistence, such that alerts are generated only when predefined thresholds remain abnormal for clinically meaningful durations rather than during brief fluctuations. Finally, the algorithm applies a multi-stage filtering strategy that adjusts alert timing according to the severity of physiological derangements, allowing more rapid notification for severe abnormalities while suppressing alerts for transient or clinically insignificant deviations. This integrated approach aims to improve the specificity of continuous monitoring and reduce alarm fatigue without compromising early detection of patient deterioration.

#### Intervention pathway

Continuous physiological data are transmitted from the bedside monitoring system to the WARD algorithm for real-time analysis. When sustained physiological abnormalities meeting predefined criteria are detected, an alert is generated and transmitted to the primary nurse through a secure handheld mobile device (Honeywell, CT30 XP, CT30PL1N, Charlotte, North Carolina), The nurse who is expected to perform an immediate bedside assessment of the patient. Based on this assessment, the primary nurse initiates the predefined clinical escalation pathway in accordance with existing hospital protocols. This may include notifying the treating physician, activating the RRT when appropriate, and implementing therapeutic interventions at the discretion of the treating team. The alert is also transmitted to the i-PROTECT command center, where it is reviewed by a dedicated critical care nurse available 24 hours a day, 7 days a week, serving as a centralized surveillance and coordination resource that supports, but does not replace, the treating clinical team. Each alert is reviewed to identify potential technical issues or non-actionable alarms before clinical notification. To ensure timely response, the intervention incorporates an automated escalation process. If an alert is not acknowledged within one minute, an electronic reminder is generated. If appropriate clinical action has not occurred within five minutes, the commands center nurse escalates the alert to the charge nurse, makes phone follow-up, and, if needed, to the treating medical team according to the predefined escalation protocol (Figure 2). All alerts, assessments, clinical actions, and patient outcomes are documented electronically within the hospital information system.

#### Data infrastructure, connectivity, and information security

The intervention depends on continuous, low-latency data transmission across three linked layers: bedside sensors, the WARD algorithmic processing engine, and the hospital’s wireless network and EMR. Reliable wireless network performance, including adequate bandwidth, redundancy, and prioritization of clinical data traffic, is a recognized prerequisite for dependable continuous ward monitoring, since intermittent connectivity or network congestion can degrade alert timeliness and reliability[31].[38] Because continuously connected monitoring devices constitute an expanded attack surface within the hospital network, the deployment incorporates network segmentation, encrypted data transmission, and access controls consistent with recommended safeguards for connected medical devices, reflecting growing regulatory and standards-body attention to cybersecurity risks associated with wirelessly networked patient monitors[32].[33] All alert, assessment, and outcome data are retained within the hospital’s governed EMR environment (BESTCare) rather than external systems, consistent with institutional data governance and privacy requirements.[24]

#### Discontinuation of the wireless monitoring

The wireless system can be discontinued if

1. NEWS-2 score drops below 5 for at least 12 hours, unless the treating team continues to have a clinical concern.
2. The patient is moved to the ICU.
3. Goals of care were changed to Do-Not-Resuscitate.
4. Treating team judgment that the wireless monitoring is no longer needed.

##### Control group

Usual care with standard intermittent monitoring but no continuous monitoring.

#### Randomization

The hospital has a total of 28 eligible wards, allowing 4 sequences of 7 wards each. Wards are randomized to receive the intervention as a 7-ward sequence each 2-month period, followed by a 1-month transition, according to a computer-generated non-stratified concealed list. The randomization list will be maintained by a research coordinator who is not involved in this trial, and the ward allocation will remain concealed from the research and clinical teams throughout the study and will be revealed for a given sequence only 1 month before the implementation of the intervention to allow training.

#### Study cohorts

The intention-to-treat (ITT) cohort includes all patients admitted to the study wards. This includes patients admitted to the intervention wards regardless of whether they are placed on the wireless monitoring system. The ITT analysis also implies that patients in the ITT cohort in the wards belonging to a particular randomization period will be analyzed in accordance with their planned randomization regardless of what happens during the trial. For example, if a ward is planned to have the monitoring system implemented during a period and for technical reasons the system is not operational, patients in that ward during that period will be analyzed as receiving the intervention. The primary analysis will be based on this population.

#### Data extraction

All data will be extracted from the electronic medical records in BESTCare. Data will be extracted as per the definitions in the attached supplementary materials.(Supplementary File 1)

#### Baseline data, physiological parameters and treatments at baseline

##### Baseline data

- Age and sex
- Admission source: emergency room, operating room, clinic, ICU, others
- Admitting ward: medical, surgical, oncology, mixed (any combination)
- Co-morbidities: end-stage renal disease, cancer without metastasis, immunocompromised non-complicated, diabetes, complicated diabetes, congestive heart failure, acquired immunodeficiency syndrome, moderate to severe chronic kidney disease, myocardial infarction, chronic pulmonary disease, peripheral vascular disease, stroke or transient ischemic attack, dementia, hemiplegia or paraplegia, connective tissue disease, peptic ulcer disease, mild liver disease, moderate to severe liver disease (Table S1 and S2)
- Charlson Comorbidity Index

###### Intervention fidelity and implementation performance

Intervention fidelity will be prospectively evaluated by reporting the proportion of patients who receive continuous wireless monitoring, duration of monitoring, alert frequency per patient-day, alert acknowledgment time, and time to initiation of clinical intervention when indicated.

##### Outcomes

###### Primary outcome

Composite endpoint of 30-day in-hospital mortality, cardiac arrest, requirement of vasopressor or intubation. Vasopressor therapy or intubation will include unplanned treatments initiated after check-in to the ward, excluding procedure-related treatments administered in operating rooms or other procedural areas.

###### Secondary outcomes

- All-cause in-hospital mortality within 30 days
- Cardiac arrest within 30 days
- Requirement of vasopressor within 30 days
- Requirement of intubation within 30 days
- Hospital length of stay (LOS), censored at 90 days
- Transfer to ICU within 30 days
- ICU-free days in the first 30 days
- RRT activation within 30 days

##### Sample size

The sample size for this stepped-wedge cluster-randomized design was calculated for 28 clusters with 5 periods (including one baseline period) using Power Analysis and Sample Size (PASS) software (PASS 15 Power Analysis and Sample Size Software (2017). NCSS, LLC. Kaysville, Utah, USA, ncss.com/software/pass) (**Table S3**). We estimated a baseline primary outcome of 6.7% based on data from the SCREEN trial,[28] and with the assumption that the intervention will reduce the primary outcome by 2%, i.e. from 6.7% to 4.7%, a sample size of 13160 subjects yields power of 0.88 and 0.91 using two-sided Wald Z-Test and significance level of 5%, and an intra-cluster correlation (ICC, a measure of the relatedness of cluster) of 0.1 and 0.2, respectively. There is no planned interim analysis, due to the nature and design of the study intervention.

##### Statistical analysis

For this trial, the analysis will be implemented under the guiding principles of the ICH E9 for the analysis of RCTs.[34] The stepped wedge design is essentially a matched design with before and after comparisons for each unit of randomization (or cluster), which is the ward in this study. Characteristics of the individuals will be summarized and compared by exposure status (intervention vs. control). All variables will be summarized and reported for the study using descriptive statistics. Interval variables will be summarized and reported in terms of n, mean and median, standard deviation, and interquartile range. Categorical variables will be summarized and reported in terms of frequency distribution. A detailed statistical analysis plan is outlined in the Online Supplement.(Supplementary File 2)

###### Analysis of the primary endpoint

The analysis for the primary outcome will be conducted on the ITT cohort. The primary outcome will be compared between the intervention group and control group at the individual level using a mixed-effects logistic regression model with the jackknife method to estimate standard errors and account for clustering within clusters.[35, 36] We will use a generalized linear mixed model with a binary distribution and a log-link function to estimate the relative risk as a measure of effect. We will include two levels of random effects to account for nested clustering within wards and periods. The best-fitting covariance structure will be selected as the one that produces the lowest Bayesian Information Criterion (BIC) value among the following candidate structures: first order of autocorrelation covariance structure, unstructured covariance structure, Toeplitz covariance structure and variance component structure (VC). The random coefficients will be modeled using G-side random effects to obtain the subject-specific estimates by defining the appropriate variance covariance structure.

###### Analysis of other outcomes

The categorical outcomes will be summarized for the intervention and control groups using numbers and percentages and analyzed in a similar manner as done for the primary outcome. The continuous outcomes will be summarized using median and interquartile range and will be analyzed using a mixed-effects Poisson model. We will include two levels of random effects to account for nested clustering within wards and periods.

###### Subgroup analysis

We will analyze the primary outcome across predefined subgroups using the same model of the primary analysis. The predefined subgroups include age ≤ 65 years and > 65 years, patients admitted to medical, surgical, and oncology wards, and patients with chronic cardiopulmonary disease versus not (as specified in Table S2: Myocardial infarction, Congestive heart failure and peripheral vascular disease). The results of the test of interaction will be reported. All analyses will be conducted with SAS version 9.4.

### Ethics and dissemination

The trial was approved by the Institutional review board (IRB) of Ministry of National Guard Health Affairs (IRB/2623/23). It is part of a quality improvement project that is implemented in the participating hospitals in phases. The stepped-wedge design is well suited to this type of pragmatic study, [37] as it allows all participating wards to eventually receive an intervention that is expected to be beneficial while enabling rigorous evaluation of its effectiveness. The study involves no invasive research procedures, and all patients receive, at a minimum, the established standard of care for clinical deterioration monitoring using NEWS2, with additional monitoring provided during the intervention periods. Moreover, the study does not interfere with or restrict routine clinical management, and all treatment decisions remain at the discretion of the treating clinical teams. Study data are obtained primarily from routinely collected electronic medical records across a large population of hospitalized ward patients. Given the pragmatic nature of the intervention, the absence of additional invasive procedures, the maintenance of standard clinical monitoring and management, and the use of routinely collected data, the study was considered minimal risk. Accordingly, the Institutional Review Board waived the requirement for individual written informed consent, consistent with the approach used in similar pragmatic studies. .[38, 39] The trial will be completed in accordance with the principles of the Ottawa statement on ethical design and conduct of cluster randomized trials.[40] The results of the i-PROTECT trial will be disseminated regardless of the direction or magnitude of effect, in accordance with the SPIRIT and CONSORT reporting guidelines. The results will be presented at relevant national and international critical care conferences. The primary results manuscript will be submitted to a peer-reviewed journal after completion and will report the primary and secondary outcomes as prespecified in the statistical analysis plan, together with a full account of any protocol deviations. Authorship will follow International Committee of Medical Journal Editors (ICMJE) criteria. No contractual agreements will limit the investigators’ access to the final trial dataset or their ability to publish independently of the funder or sponsor.

## Discussion

Clinical deterioration on hospital wards remains a major cause of preventable morbidity and mortality despite widespread implementation of rapid response systems and early warning scores. Although intermittent physiological observations combined with NEWS-2 and RRT. have improved recognition of deteriorating patients, clinically significant abnormalities frequently occur between scheduled assessments, potentially delaying intervention. Continuous wireless physiological monitoring has emerged as a promising strategy to bridge this surveillance gap; however, existing evidence has been limited largely to feasibility studies, before-and-after evaluations, and relatively small randomized trials, leaving uncertainty regarding its impact on patient-centered clinical outcomes.

The i-PROTECT trial has been designed to address this important evidence gap by evaluating continuous wireless monitoring within a real-world hospital environment using a stepped-wedge cluster randomized design. Unlike previous studies that primarily evaluated the monitoring device itself, this intervention represents a comprehensive care pathway integrating continuous physiological monitoring, intelligent algorithmic analysis, centralized expert surveillance, structured communication, predefined escalation protocols, and standardized electronic documentation. The intervention therefore aims not only to detect physiological abnormalities earlier but also to ensure that early detection translates into timely clinical assessment and appropriate intervention, which ultimately determine patient outcomes. If effective, this intervention could provide a scalable framework for integrating continuous physiological monitoring into existing rapid response systems. Because the intervention leverages existing nursing workflows and electronic medical records rather than replacing them, implementation may be feasible in other hospitals with mature rapid response systems while allowing adaptation to local organizational structures. From an information technology perspective, successful scale-up of continuous ward monitoring programs elsewhere has depended on phased, system-wide implementation planning, interoperable integration with the EMR, and sustained 24/7 technical and network support rather than on the monitoring device alone.[41] This suggests that the transferability of the i-PROTECT pathway to other institutions will depend not only on clinical and organizational readiness but also on the maturity of the receiving hospital’s network infrastructure, EMR interoperability capabilities, and IT governance structures.

Several features distinguish the present study from previous investigations. First, the intervention is implemented within an established healthcare system that already incorporates NEWS-2 scoring, an experienced RRT, and an integrated electronic medical record. Consequently, the trial evaluates the incremental benefit of continuous monitoring when added to contemporary standard care. Second, the study targets patients at increased risk of deterioration, including patients with NEWS-2 scores of 5 or greater, patients with clinician concern despite lower NEWS-2 scores, post-ICU patients, and patients following RRT activation. This risk-enriched approach is expected to improve monitoring efficiency while minimizing unnecessary monitoring of low-risk patients. Third, the intervention incorporates advanced signal-processing algorithms that prioritize persistent and clinically meaningful physiological abnormalities while suppressing transient artifacts, addressing one of the major barriers to implementing continuous monitoring: alarm fatigue.

The choice of a stepped-wedge cluster randomized design reflects both methodological and practical considerations. The intervention is implemented at the ward level and involves substantial workflow modifications, staff education, centralized monitoring, and organizational changes that preclude individual patient randomization without substantial contamination. The stepped-wedge design permits sequential implementation across hospital wards while ensuring that all participating wards eventually receive the intervention, an approach that is both operationally feasible and ethically acceptable for evaluating a quality improvement initiative expected to provide benefit. Furthermore, this design allows adjustment for temporal trends while preserving the advantages of randomized allocation.

The primary outcome was selected to capture clinically meaningful manifestations of severe physiological deterioration that are most likely to be modified by earlier recognition and intervention. The composite endpoint includes 30-day in-hospital mortality, cardiac arrest, initiation of vasopressor therapy, and endotracheal intubation. Mortality and cardiac arrest represent patient-centered outcomes, whereas vasopressor therapy and endotracheal intubation reflect progression to cardiovascular or respiratory failure requiring major life-sustaining interventions. Combining these outcomes improves statistical efficiency while capturing the continuum of clinically important deterioration that continuous monitoring is intended to prevent. Importantly, outcomes such as ICU admission, unexpected ICU transfer, and RRT activation were intentionally excluded from the primary endpoint because they are process measures substantially influenced by institutional practices, clinician judgment, bed availability, and local escalation protocols. Moreover, successful implementation of continuous monitoring may appropriately increase early ICU transfer or RRT activation by facilitating earlier recognition of clinical deterioration, making these outcomes difficult to interpret as indicators of treatment benefit. Accordingly, these variables will be evaluated as secondary outcomes that provide insight into the mechanisms through which the intervention influences patient outcomes, while the primary composite remains focused on objective clinical events that directly reflect failure of physiological compensation.

This study has several strengths. The pragmatic design reflects routine clinical practice and therefore enhances external validity. The intervention leverages existing hospital infrastructure, including the electronic medical record, established escalation pathways, and rapid response systems, thereby facilitating implementation and future scalability. In addition, the large sample size, cluster randomization, standardized electronic data collection, and centralized monitoring are expected to provide robust estimates of clinical effectiveness while minimizing selection bias and missing outcome data.

Several limitations should also be acknowledged. Blinding of healthcare providers is not feasible because of the nature of the intervention. As with all stepped-wedge trials, secular changes in clinical practice over time may influence outcomes, although these will be accounted for in the statistical analysis. Some degree of contamination may occur because healthcare professionals may rotate between intervention and control wards and may alter their clinical practice following exposure to the intervention. Furthermore, the study is conducted within a single tertiary academic healthcare system with mature rapid response infrastructure and a comprehensive electronic medical record; therefore, generalizability to hospitals with different organizational structures or fewer resources may be limited. The intervention combines multiple components (continuous monitoring, algorithmic analysis, centralized review, and structured escalation), making it difficult to determine the individual contribution of each component to the overall treatment effect.

If successful, the i-PROTECT trial will provide high-quality randomized evidence regarding the effectiveness of integrating continuous wireless monitoring into routine ward care. The findings are expected to inform future implementation strategies, clinical practice guidelines, and healthcare policy regarding continuous monitoring of high-risk ward patients while providing an evidence-based framework for integrating digital monitoring technologies into modern inpatient care.

### Trial status

The trial started on Feb 1, 2026, and is expected to complete patient follow-up by the end of May 2027.

## Data Availability

The data supporting this study's findings are available from the corresponding author upon reasonable request as per the regulations of King Abdullah International Medical Research Center (KAIMRC).

## Declaration

### Author contributions

YA is the Chief Investigator responsible for the conception, design and development of the protocol, analytical plan, drafting and critical revision of the manuscript for important and intellectual content.

AA, TD, AK, KA, HD, NT, RH, AM, AC, KK, NA, AA, SA, SYQ, SQ, AR, ZY, AH, RA, WM, MK, MM, GH, RA, MS, KS, AE, WR, AS, and AD contributed to the development of the protocol and critical revision of the manuscript for important intellectual content.

HAr and BY provided the statistical expertise for the statistical plan. All authors read and approved the final manuscript

### Ethics approval and consent to participate

The study has been approved by the Institutional Review Board (IRB) of the Ministry of National Guard Health Affairs (MNGHA) (IRB/2623/23). Due to the pragmatic nature of the study, Institutional Review Board waived the requirement for individual written informed consent

### Consent for publication

Not applicable

### Patient and public involvement

Patients and/or the public were not involved in the design, or conduct, or reporting, or dissemination plans of this research.

### Provenance and peer review

Not commissioned; externally peer-reviewed.

## References

1. Jones D, Mitchell I, Hillman K, Story D: Defining clinical deterioration. Resuscitation 2013, 84(8):1029–1034.

2. Buist M, Bernard S, Nguyen TV, Moore G, Anderson J: Association between clinically abnormal observations and subsequent in-hospital mortality: a prospective study. Resuscitation 2004, 62(2):137–141.

3. Al-Qahtani S, Al-Dorzi HM, Tamim HM, Hussain S, Fong L, Taher S, Al-Knawy BA, Arabi Y: Impact of an intensivist-led multidisciplinary extended rapid response team on hospital-wide cardiopulmonary arrests and mortality. Critical care medicine 2013, 41(2):506–517.

4. Arabi YM, Al-Dorzi HM, Alamry A, Hijazi Re, Alsolamy S, Al Salamah M, Tamim HM, Al-Qahtani S, Al-Dawood A, Marini AM: The impact of a multifaceted intervention including sepsis electronic alert system and sepsis response team on the outcomes of patients with sepsis and septic shock. Annals of intensive care 2017, 7(1):1–10.

5. Hogan H, Healey F, Neale G, Thomson R, Vincent C, Black N: Preventable deaths due to problems in care in English acute hospitals: a retrospective case record review study. BMJ Quality & Safety 2012, 21(9):737–745.

6. Smith GB: In-hospital cardiac arrest: is it time for an in-hospital ‘chain of prevention’? Resuscitation 2010, 81(9):1209–1211.

7. Armitage M, Eddleston J, Stokes T: Recognising and responding to acute illness in adults in hospital: summary of NICE guidance. Bmj 2007, 335(7613):258–259.

8. Gonem S, Draicchio D, Mohamed A, Wood S, Shiel K, Briggs S, McKeever TM, Shaw D: Physiological deterioration prior to in-hospital cardiac arrest: What does the National Early Warning Score-2 miss? Resusc Plus 2024, 20:100788.

9. Weenk M, van Goor H, Frietman B, Engelen LJ, van Laarhoven CJ, Smit J, Bredie SJ, van de Belt TH: Continuous monitoring of vital signs using wearable devices on the general ward: pilot study. JMIR mHealth and uHealth 2017, 5(7):e91.

10. Breteler MJ, Huizinga E, van Loon K, Leenen LP, Dohmen DA, Kalkman CJ, Blokhuis TJ: Reliability of wireless monitoring using a wearable patch sensor in high-risk surgical patients at a step-down unit in the Netherlands: a clinical validation study. BMJ open 2018, 8(2):e020162.

11. Breteler MJ, KleinJan EJ, Dohmen DA, Leenen LP, van Hillegersberg R, Ruurda JP, van Loon K, Blokhuis TJ, Kalkman CJ: Vital signs monitoring with wearable sensors in high-risk surgical patients: a clinical validation study. Anesthesiology 2020, 132(3):424–439.

12. Kjaergaard K, Molgaard J, Rasmussen SM, Meyhoff CS, Aasvang EK: The effect of technical filtering and clinical criteria on alert rates from continuous vital sign monitoring in the general ward. Hosp Pract (1995) 2023, 51(5):295–302.

13. Bellomo R, Ackerman M, Bailey M, Beale R, Clancy G, Danesh V, Hvarfner A, Jimenez E, Konrad D, Lecardo M: A Controlled Trial of Electronic Automated Advisory Vital Signs Monitoring in General Hospital Wards. Critical care medicine 2012, 40(8):2349–2361.

14. Subbe CP, Duller B, Bellomo R: Effect of an automated notification system for deteriorating ward patients on clinical outcomes. Critical Care 2017, 21(1):1–9.

15. Taenzer AH, Pyke JB, McGrath SP, Blike GT: Impact of pulse oximetry surveillance on rescue events and intensive care unit transfers: a before-and-after concurrence study. The Journal of the American Society of Anesthesiologists 2010, 112(2):282–287.

16. Brown H, Terrence J, Vasquez P, Bates DW, Zimlichman E: Continuous monitoring in an inpatient medical-surgical unit: a controlled clinical trial. The American journal of medicine 2014, 127(3):226–232.

17. Rowland B, Saha A, Motamedi V, Bundy R, Winsor S, McNavish D, Lippert W, Khanna AK: Impact on Patient Outcomes of Continuous Vital Sign Monitoring on Medical Wards: Propensity-Matched Analysis. J Med Internet Res 2025, 27:e66347.

18. Downey C, Randell R, Brown J, Jayne DG: Continuous versus intermittent vital signs monitoring using a wearable, wireless patch in patients admitted to surgical wards: pilot cluster randomized controlled trial. Journal of medical Internet research 2018, 20(12):e10802.

19. Khanna AK, O’Connell NS, Saha AK, Hicks MH, Weller RS, Harris L, Cusson BD, Faris A, Huffman CS, Segal S et al: Continuous vs Intermittent Postoperative Vital Sign Monitoring: A Cluster Randomized Crossover Trial. JAMA Netw Open 2026, 9(3):e263290.

20. Molgaard J, Gronbaek KK, Rasmussen SS, Eiberg JP, Jorgensen LN, Achiam MP, Rohrsted M, Singh UM, Hoang TH, Sogaard M et al: Continuous Vital Sign Monitoring at the Surgical Ward for Improved Outcomes After Major Noncardiac Surgery: A Randomized Clinical Trial. Anesth Analg 2025, 141(4):807–817.

21. Chan AW, Tetzlaff JM, Altman DG, Laupacis A, Gotzsche PC, Krleza-Jeric K, Hrobjartsson A, Mann H, Dickersin K, Berlin JA et al: SPIRIT 2013 statement: defining standard protocol items for clinical trials. Ann Intern Med 2013, 158(3):200–207.

22. Hemming K, Taljaard M, McKenzie JE, Hooper R, Copas A, Thompson JA, Dixon-Woods M, Aldcroft A, Doussau A, Grayling M et al: Reporting of stepped wedge cluster randomised trials: extension of the CONSORT 2010 statement with explanation and elaboration. BMJ 2018, 363:k1614.

23. Hoffmann TC, Glasziou PP, Boutron I, Milne R, Perera R, Moher D, Altman DG, Barbour V, Macdonald H, Johnston M et al: Better reporting of interventions: template for intervention description and replication (TIDieR) checklist and guide. BMJ 2014, 348:g1687.

24. Arabi YM, Al Ghamdi AA, Al-Moamary M, Al Mutrafy A, AlHazme RH, Al Knawy BA: Electronic medical record implementation in a large healthcare system from a leadership perspective. BMC Med Inform Decis Mak 2022, 22(1):66.

25. Avakian A: Closing the Loop: A Software-Based Middleware Framework for Automated Vital Sign Integration With Cloud-Based Electronic Medical Records (EMRs). Cureus 2025, 17(8):e90513.

26. HL7 International. Caliper Accelerator: Medical Device Interoperability Implementation Community. .

27. Royal College of Physicians. National Early Warning Score (NEWS) 2. https://www.rcp.ac.uk/resources/national-early-warning-score-news-2/. Accessed Aug 31-2026. .

28. Arabi YM, Alsaawi A, Alzahrani M, Al Khathaami AM, AlHazme RH, Al Mutrafy A, Al Qarni A, Vishwakarma RK, Al Anazi R, Al Qasim E et al: Electronic Sepsis Screening Among Patients Admitted to Hospital Wards: A Stepped-Wedge Cluster Randomized Trial. JAMA 2024.

29. Masimo. https://www.masimo.com/products/root/. Accessed Aug 31–2026.

30. Ward 24/7. www.ward247.com. Accessed on Aug 31, 2026.

31. 7SIGNAL. Wi-Fi for Healthcare and Hospitals: Optimization Considerations for Clinical Connectivity. . In.

32. IEEE Standards Association. Medical Device Security: Data Risks for Connected Medical Devices. .

33. Menon V: Cybersecurity breaches in medical devices: analyzing FDA safety communications in response to patient security concerns. Front Digit Health 2026, 8:1701551.

34. International Council for Harmonisation of Technical Requirements for Pharmaceuticals for Human Use (ICH). E9 Statistical Principles for Clinical Trials. London: ICH; 1998. Available at: https://www.ich.org/page/efficacy-guidelines.

35. Hussey MA, Hughes JP: Design and analysis of stepped wedge cluster randomized trials. Contemp Clin Trials 2007, 28(2):182–191.

36. Shakeshaft A, Doran C, Petrie D, Breen C, Havard A, Abudeen A, Harwood E, Clifford A, D’Este C, Gilmour S et al: The effectiveness of community action in reducing risky alcohol consumption and harm: a cluster randomised controlled trial. PLoS Med 2014, 11(3):e1001617.

37. Mdege ND, Man MS, Taylor Nee Brown CA, Torgerson DJ: Systematic review of stepped wedge cluster randomized trials shows that design is particularly used to evaluate interventions during routine implementation. J Clin Epidemiol 2011, 64(9):936–948.

38. Gonzales R, Anderer T, McCulloch CE, Maselli JH, Bloom FJ, Graf TR, Stahl M, Yefko M, Molecavage J, Metlay JP: A Cluster-Randomized Trial of Decision Support Strategies for Reducing Antibiotic Use for Acute Bronchitis. JAMA internal medicine 2013, 173(4):267–273.

39. van Wyk JT, van Wijk MA, Sturkenboom MC, Mosseveld M, Moorman PW, van der Lei J: Electronic alerts versus on-demand decision support to improve dyslipidemia treatment: a cluster randomized controlled trial. Circulation 2008, 117(3):371–378.

40. Weijer C, Grimshaw JM, Eccles MP, McRae AD, White A, Brehaut JC, Taljaard M, Ottawa Ethics of Cluster Randomized Trials Consensus G: The Ottawa Statement on the Ethical Design and Conduct of Cluster Randomized Trials. PLoS Med 2012, 9(11):e1001346.

41. Nguyen NA, Lee G, Holderread B, Holman T, Pletcher S, Schwartz R: Scaling Wireless Continuous Vital Sign Monitoring Across an 8-Hospital Health System: Digital Health Implementation Report. JMIR Med Inform 2026, 14:e78216.

