## Supplementary File 1 for "Wireless Monitoring of Patients Admitted to Hospital Wards: A Stepped-Wedge Cluster Randomized Trial (i-PROTECT Trial): Clinical Study Protocol and Statistical Analysis Plan"

**Table S1:** Definitions used in the i-PROTECT trial.

| Item | Description |
|---|---|
| **Time 0** | Date and time when the patient is checked in for the first time to the ward. If transferred to different wards, take the first date and time only. |
| **Alert time** | When the patient meets the alert criteria; for patients who have multiple alerts, we will take only the first alert |
| **Observation time** | The whole hospital stay from time of check-in to the ward to time of discharge from the hospital censored at 30 days. The observation period excludes stays in the operating room, daycare operation room, operation cardiac, outpatient, post-anesthesia care unit, dental outpatient, day surgery, daycare outpatient, intensive care unit, trauma intensive care unit, surgical intensive care unit, Intermediate care unit, progressive care unit, medical cardiac intensive care unit, neurointensive care unit, burn unit). |
| **Alert population** | Patients who had one alert or more during hospitalization. However, detailed data will be included only for the first alert. For subsequent alerts, only alert count will be provided. |

**Table S2:** The ICD-10-AM codes for comorbid conditions.

| Comorbidity | ICD-10-AM Codes |
|---|---|
| Myocardial infarction | I21.x, I22.x, I25.2 |
| Congestive heart failure | I09.9, I11.0, I13.0, I13.2, I25.5, I42.0, I42.5 - I42.9, I43.x, I50.x, P29.0 |
| Peripheral vascular disease | I70.x, I71.x, I73.1, I73.8, I73.9, I77.1, I79.0, I79.2, K55.1, K55.8, K55.9, Z95.8, Z95.9 |
| Cerebrovascular disease | G45.x, G46.x, H34.0, I60.x - I69.x |
| Dementia | F00.x - F03.x, F05.1, G30.x, G31.1 |
| Chronic pulmonary disease | I27.8, I27.9, J40.x - J47.x, J60.x - J67.x, J68.4, J70.1, J70.3 |
| Rheumatic disease | M05.x, M06.x, M31.5, M32.x - M34.x, M35.1, M35.3, M36.0 |
| Peptic ulcer disease | K25.x - K28.x |
| Mild liver disease | B18.x, K70.0 - K70.3, K70.9, K71.3 - K71.5, K71.7, K73.x, K74.x, K76.0, K76.2 - K76.4, K76.8, K76.9, Z94.4 |
| Diabetes without chronic complication | E10.0, E10.1, E10.6, E10.8, E10.9, E11.0, E11.1, E11.6, E11.8, E11.9, E12.0, E12.1, E12.6, E12.8, E12.9, E13.0, E13.1, E13.6, E13.8, E13.9, E14.0, E14.1, E14.6, E14.8, E14.9 |
| Diabetes with chronic complication | E10.2 - E10.5, E10.7, E11.2 - E11.5, E11.7, E12.2 - E12.5, E12.7, E13.2 - E13.5, E13.7, E14.2 - E14.5, E14.7 |
| Hemiplegia or paraplegia | G04.1, G11.4, G80.1, G80.2, G81.x, G82.x, G83.0 - G83.4, G83.9 |
| Renal disease | I12.0, I13.1, N03.2 - N03.7, N05.2 - N05.7, N18.x, N19.x, N25.0, Z49.0 - Z49.2, Z94.0, Z99.2 |
| Any malignancy, including lymphoma and leukemia, except malignant neoplasm of skin | C00.x - C26.x, C30.x - C34.x, C37.x - C41.x, C43.x, C45.x - C58.x, C60.x - C76.x, C81.x - C85.x, C88.x, C90.x - C97.x |
| Moderate or severe liver disease | I85.0, I85.9, I86.4, I98.2, K70.4, K71.1, K72.1, K72.9, K76.5, K76.6, K76.7 |
| Metastatic solid tumor | C77.x - C80.x |
| AIDS/HIV | B20.x - B22.x, B24.x |

**Table S3:** Sample size calculation for the stepped wedge cluster design.

** = two months duration time period with one month transition period

| Power | Design S/T/R | Cluster Clusters K | Sample Size M/m | Trt Size N | Ctrl Prop P1 | Prop P2 | Diff D1 | ICC | Alpha | Duration |
|---|---|---|---|---|---|---|---|---|---|---|
| 0.88279 | 4/5/07 | 28 | 470/94 | 13160 | 0.067 | 0.047 | 0.02 | 0.1 | 0.05 | 15.00 |
| 0.91445 | 4/5/07 | 28 | 470/94 | 13160 | 0.067 | 0.047 | 0.02 | 0.2 | 0.05 | 15.00 |

** = two months duration time period with one month transition period
* **=three months duration time period with one month transition period
* ***=three and a half months duration time period with one month transition period
Report Definitions
Power is the probability of rejecting the null hypothesis when the alternative is true. It should be close to one.
S is the number of steps in the study design. S = T - 1.
T is the number of time periods in the study, including the baseline. T = S + 1
R is the number clusters switching from control to treatment at each step.
K is the total number of clusters to be randomized. K = S × R.
M is the average number of subjects per cluster. M = m × T.
m is the average number of subjects per cluster per time period. M = m × T.
N is total sample size from all clusters and time periods combined. m = M/T.
P1 is the treatment proportion, assuming the alternative hypothesis.
P2 is the control, standard, reference, or baseline proportion.
D1 = P1 - P2 is the difference assuming the alternative hypothesis (H1).
ICC is the intracluster correlation coefficient.
Alpha is the probability of rejecting the null hypothesis when it is true.
