## Supplementary File 2 for "Wireless Monitoring of Patients Admitted to Hospital Wards: A Stepped-Wedge Cluster Randomized Trial (i-PROTECT Trial): Clinical Study Protocol and Statistical Analysis Plan"

**Trial registration:** ClinicalTrials.gov Identifier: **NCT06644599**.

### Background

Continuous wireless physiological monitoring has the potential to improve the early recognition of clinical deterioration in hospitalized ward patients. However, robust evidence from large randomized trials demonstrating that integrated monitoring systems improve clinically important patient outcomes remains limited. The Integrated Proactive Wireless Monitoring for Clinical Deterioration (i-PROTECT) Trial was designed to address this evidence gap. Using a pragmatic stepped-wedge cluster randomized design, the trial evaluates whether an integrated care pathway combining continuous wireless physiological monitoring, algorithm-guided alerting, centralized clinical surveillance, and structured escalation reduces cardiopulmonary failure and 30-day in-hospital mortality compared with standard intermittent monitoring. We hypothesize that this comprehensive monitoring strategy will facilitate earlier recognition of clinical deterioration and improve patient outcomes.

The purpose of this statistical analysis plan (SAP) is to prospectively define the statistical methods for the iPROTECT trial before database lock and before inspection of comparative outcome results. The SAP is based on the prespecified iPROTECT supplementary tables and follows the structure and level of detail of the published SCREEN SAP.

The SAP will be implemented according to the intention-to-treat principle and the International Council for Harmonisation E9 statistical principles.(1) Reporting will follow CONSORT 2010, the CONSORT extension for cluster randomized trials, and the CONSORT extension for stepped-wedge cluster randomized trials.(2-4)

All deviations from this SAP will be documented, dated, justified, and identified as prespecified or post hoc in the final manuscript.

### Methods

#### Study design

iPROTECT is a pragmatic, single-center, stepped-wedge cluster randomized trial. The hospital ward is the unit of randomization. Twenty-eight wards are allocated to four sequences, with seven wards in each sequence. All wards begin in the control condition and sequentially cross to the intervention condition according to the randomized implementation schedule.

The intervention is an integrated care pathway consisting of continuous wireless physiological monitoring, algorithmic alert processing, notification of the primary nurse on a hand-held device, centralized 24-hour review by the iPROTECT command-center nurse, structured escalation, and electronic documentation.

The control condition is usual ward care with intermittent vital-sign monitoring and NEWS-2-based escalation.

The planned sample size is 13,160 ward admissions.

**Cluster, sequence, period, and step definitions:** The unit of randomization (cluster) is the hospital ward. Wards initially receive the control condition (usual care without continuous wireless monitoring) and subsequently cross over to the intervention condition (the integrated iPROTECT monitoring pathway) according to a randomized stepped-wedge design. The wards are allocated to four sequences, each comprising seven wards, with crossover occurring in four sequential steps. The study consists of five analysis periods, each lasting two months. A one-month transition period is incorporated between consecutive periods to allow implementation, staff training, and stabilization of the intervention before outcome evaluation. (Table S1)

**Randomization and concealment**

The order in which ward sequences cross from control to intervention is determined by a computer-generated, non-stratified concealed randomization list. The allocation for each sequence is released approximately one month before implementation to permit staff preparation and training. No cluster is expected to revert from intervention to control.

The randomization schedule, ward roster, date and time of crossover, and any implementation delays will be retained in the trial master file.

**Primary analysis:** The primary analysis targets the effect of assignment to the iPROTECT intervention compared with usual care among all eligible patients admitted to participating wards during randomized control or intervention periods. The primary outcome is a composite of 30-day in-hospital mortality, cardiac arrest, initiation of vasopressor therapy, or endotracheal intubation. The primary analysis will follow the intention-to-treat principle, whereby patients will be analyzed according to the randomized assignment of their ward regardless of actual receipt or duration of continuous monitoring. Intercurrent events, including ward transfers, temporary device or system failures, nonadherence to the intervention, ICU transfer, and treatment decisions made by the clinical team, will be handled using a treatment-policy strategy. The treatment effect will be expressed as the relative risk with corresponding 95% confidence intervals, with the absolute risk difference and number needed to treat reported as supplementary measures where estimable.

**Primary objective**

To determine whether assignment of a hospital ward to the iPROTECT intervention reduces the risk of cardiopulmonary failure within 30 days of ward admission compared with assignment to usual care.

**Secondary objectives**

- To estimate the effect of the intervention on each component of the primary composite.
- To estimate the effect on ICU transfer, CCRT activation, hospital length of stay, and ICU-free days.
- To characterize intervention reach, alert burden, acknowledgment, response, monitoring duration, and technical performance.
- To evaluate consistency of the primary treatment effect across prespecified subgroups.

### ANALYSIS POPULATIONS

#### INTENTION-TO-TREAT COHORT

The primary analysis population will include all eligible patients checked in to eligible wards during randomized analysis periods. Patients will be analyzed according to the condition assigned to the ward and period, regardless of actual device placement, duration of monitoring, technical interruptions, or adherence to the intervention pathway.
A patient transferred between study wards during one hospitalization will be assigned to the first eligible study ward for the primary analysis.

#### Monitored cohort

The monitored cohort will include patients in intervention wards who were connected to the iPROTECT monitoring system. This cohort will be used for descriptive implementation, alert, and fidelity analyses and will not replace the ITT cohort for causal effectiveness analyses.

### GENERAL STATISTICAL PRINCIPLES

All statistical tests will be two-sided, with a significance level of 0.05 for the primary outcome. Treatment effects will be reported as point estimates with corresponding 95% confidence intervals. Because there is a single primary outcome, no adjustment for multiplicity will be applied to the primary analysis. Secondary outcomes and subgroup analyses will be considered supportive and interpreted using the Benjamini–Hochberg false discovery rate procedure, as described in the Multiplicity section. Baseline demographic and clinical characteristics will be summarized descriptively by randomized treatment group without formal statistical comparisons. Categorical variables will be presented as frequencies and percentages, while continuous variables will be summarized as mean (standard deviation) for approximately normally distributed data or median (first and third quartiles) for skewed distributions. All comparative analyses will account for the stepped-wedge cluster randomized design by incorporating calendar period and ward-level clustering in the prespecified statistical models. The treatment effects will be reported using the prespecified effect measure for the different outcomes, indicating relative risks and risk differences for binary outcomes and regression coefficients for continuous outcomes when applicable. Analyses will be performed only after completion of data cleaning, adjudication of derived outcomes, reconciliation of ward and period assignments, and formal database lock. Statistical analyses will be conducted using SAS and R statistical software, with all programming performed according to predefined quality-control procedures.

### DATA PREPARATION AND DERIVATIONS

#### Index date and time origin

The index date for the ITT analysis will be the date and time of check-in to the eligible study ward (Table S1). Outcomes will be assessed from that time through the earliest of 30 days, hospital discharge, or death, as applicable. Readmissions will be treated as separate index admissions unless the protocol specifies otherwise.

**Treatment assignment**

Treatment assignment will be derived from ward, calendar date/time, randomized sequence, and implementation status. Transition months will be coded separately. The analysis will exclude patients admitted during transition months.

**Baseline variables**

Baseline variables will include age, sex, admission source, ward type, comorbidities, Charlson Comorbidity Index, post-ICU status, CCRT activation status, and NEWS-2 score of 5 or higher. Comorbidities will be derived from documented diagnoses using the prespecified ICD-10 coding algorithms.

**Primary composite outcome**

The primary composite will be coded as present if at least one of the following occurs within 30 days of the index ward admission and before hospital discharge:

- All-cause in-hospital death.
- Code blue activation.
- Initiation of vasopressor therapy.
- Endotracheal intubation and invasive mechanical ventilation.

For patients with more than one component, the composite will be counted once. Component dates and the first qualifying component will be retained. Elective intubation and vasopressors administered in the operating room, procedural areas, or during elective anesthesia are excluded.

**Secondary outcome definitions**

Secondary outcomes include 30-day in-hospital mortality, defined as death from any cause during the index hospitalization within 30 days of admission to the study ward; code blue activation, defined as the first qualifying code blue event within 30 days; new nonelective initiation of protocol-defined systemic vasopressor therapy within 30 days; new nonelective endotracheal intubation within 30 days; transfer to an intensive care unit (ICU) within 30 days after index ward admission; and at least one Critical Care Response Team (CCRT) activation within 30 days. Hospital length of stay will be calculated as the number of days from hospital admission to discharge or death and will be censored at 90 days. ICU-free days will be defined as the number of days alive and not admitted to the ICU during the first 30 days after index ward admission, with patients who die before day 30 assigned a value of zero ICU-free days.

### DESCRIPTIVE AND IMPLEMENTATION ANALYSES

**Participant and cluster flow**

A CONSORT flow diagram adapted for stepped-wedge cluster randomized trials will present wards by sequence and period, number of admissions, exclusions, transition-period records, intervention exposure, and records included in each analysis population. (4)

For each period and sequence, the number of active clusters, control clusters, mean cluster size and variance will be reported (Figure S1).

**Baseline characteristics**

Baseline characteristics will be presented by randomized exposure condition for the ITT cohort using Table S2. No hypothesis tests will be performed for baseline imbalance.

Intervention reach and fidelity

We will report:

- Number and percentage of eligible patients who received continuous monitoring overall and within each eligibility category.
- Duration of monitoring.
- Number and percentage of monitored patients with at least one alert.
- Alert count per patient and per monitored patient-day.
- Time from monitoring initiation to first alert.
- Alert type, including respiratory, oxygen saturation, heart rate, blood pressure, and other prespecified categories.
- Percentage of alerts acknowledged by the nurse.
- Time from alert generation to acknowledgment, summarized continuously and in prespecified categories.
- Action taken.
- False, technical, and clinically actionable alert rates.

**PRIMARY OUTCOME ANALYSIS**

**Primary model**

The primary composite outcome will be compared between intervention and control conditions at the patient level using a generalized linear mixed model with a binomial distribution and log link to estimate the relative risk.(5, 6) The model will include:

- A fixed effect for treatment condition.
- Fixed effects for calendar period to account for secular trends.
- A random intercept for ward to account for within-ward correlation. The intracluster correlation coefficient (ICC) will be estimated from the fitted model to quantify within-ward correlation and will be reported, where applicable.

If supported by the final design and software implementation, a random ward-by-period effect or correlation structure for repeated ward-period observations will be considered. The covariance structure will be prespecified before unblinding and selected from plausible structures using convergence, parsimony, and Bayesian Information Criterion, without reference to the treatment estimate. Models inherently accommodate unequal cluster sizes; no weighting will be applied.

Robust or small-sample corrected standard errors will be used because there are 28 clusters. The preferred small-sample correction will be the Kenward-Roger or between-within correction where compatible with the selected model. A jackknife variance estimator may be used to maintain consistency with the SCREEN SAP if technically supported.

The primary effect will be reported as relative risk with 95% CI and two-sided p-value. Model-based marginal risks in each condition, absolute risk difference, and number needed to treat will also be reported where estimable. We will also estimate risk difference.(7) Model diagnostics and convergence

- Convergence status, boundary estimates, and covariance parameter estimates will be reviewed.
- Overdispersion will be assessed using the Pearson chi-square statistic divided by its degrees of freedom where applicable.
- If sparse events produce quasi-separation, penalized likelihood or Firth-type correction will be considered.
- If the log-binomial model fails to converge, a modified Poisson mixed model with robust variance will be the first fallback.
- If that model also fails, a mixed-effects logistic regression will be fitted and the odds ratio reported, with marginal standardized risks and risk ratio estimated where feasible.

**Primary analysis presentation**

The primary table will show event counts and percentages by treatment condition, relative risk, 95% CI, absolute risk difference, and p value.

### SECONDARY OUTCOME ANALYSES

#### Binary secondary outcomes

Thirty-day mortality, code blue, vasopressor initiation, intubation, ICU transfer, and CCRT activation will be analyzed using the same stepped-wedge mixed-model framework as the primary outcome. Results will be reported as relative risks with 95% CIs. These analyses will be considered secondary and interpreted alongside FDR-adjusted p values.

Continuous variables:

Hospital length of stay and ICU-free days, censored at 90 days, will be summarized using median and quartiles. Because these variables are non-negative and typically skewed, will use a mixed-effects model appropriate to the observed distribution, preferably a negative binomial or zero-inflated count model if required. A rank-based cluster-adjusted sensitivity analysis may be added if model assumptions are poor.

### SUBGROUP ANALYSES

The primary outcome will be analyzed across the following prespecified subgroups: Age 65 years or younger versus older than 65 years, Medical, surgical, oncology, and mixed wards and Chronic cardiopulmonary disease versus no chronic cardiopulmonary disease.
Subgroup treatment effects will be estimated by adding the subgroup main effect and treatment-by-subgroup interaction to the primary model. Interaction p values, not within-subgroup significance, will be used to assess heterogeneity. Results will be displayed in a forest plot. FDR-adjusted interaction p values will be presented.

### SENSITIVITY ANALYSES

Prespecified sensitivity analyses will be performed to evaluate the robustness of the primary findings. First, the primary analysis will be repeated using a covariate-adjusted model that includes age, sex, ward type, admission source, Charlson Comorbidity Index, post-ICU status, prior CCRT activation, and NEWS-2 eligibility status, provided that model convergence and data completeness are adequate. Second, if the primary log-binomial mixed-effects model fails to converge or demonstrates poor model fit, a modified Poisson mixed-effects model with robust variance estimation will be used as an alternative to estimate relative risks. Third, because there are fewer than 50 clusters, we will conduct a sensitivity analysis using a small sample correction with the Kenward-Roger method.(5, 8, 9) The consistency of results across sensitivity analyses will be evaluated by comparing the direction, magnitude, and precision of treatment effect estimates rather than relying solely on statistical significance. These analyses are intended to assess the robustness of the primary findings and will be considered supportive rather than confirmatory.

### MISSING DATA

Primary treatment assignment and major clinical outcomes are expected to be available for nearly all eligible patients through the electronic medical record (EMR). The extent, pattern, and potential mechanisms of missing data will be summarized for all study variables. No imputation will be performed for descriptive analyses. Missingness of the primary outcome is expected to be negligible because outcome data are routinely captured within the EMR. However, if missing primary outcome data exceed 1% of the study population, sensitivity analyses will be performed using plausible best-case and worst-case scenarios and, where appropriate, multiple imputation. For adjusted analyses, the mechanism of missingness for baseline covariates will be assessed and classified as missing completely at random (MCAR), missing at random (MAR), or potentially missing not at random (MNAR). Covariates considered to be missing at random will be imputed using multiple imputation by chained equations (MICE), with imputation models incorporating treatment assignment, calendar period, ward, study outcomes, and other variables predictive of missingness and the missing values themselves. The number of imputations will be at least equal to the percentage of incomplete cases, with a minimum of 20 imputations. Parameter estimates from the imputed datasets will be combined using Rubin's rules.(10) The primary analysis will be based on the complete-case dataset, with multiple imputation analyses considered supportive and used to evaluate the robustness of the findings.

Other variables used in the model adjustment model will be assessed and characterized in terms of their pattern (i.e., Missing Completely at Random, Missing at Random, Missing Not at Random). For Missing Completely at Random data, all analyses will be based on a list-wise deletion approach where observation will complete values will be only considered for analysis. For variables with values Missing at Random, multiple imputation techniques will be utilized to impute the missing values as suggested by Rubin.(10) And for variables with values Missing Not at Random, a pattern-mixture model technique will be used to impute the missing values.(11)

### MULTIPLICITY

The primary outcome will be tested without multiplicity adjustment. Secondary outcome and subgroup interaction p values will be adjusted using the Benjamini-Hochberg false discovery rate procedure with q=0.05.(12) Both nominal and FDR-adjusted p values will be reported.
Confidence intervals for secondary outcomes will remain 95% descriptive intervals 14.

#### Protocol deviations

Protocol deviations will be prospectively documented and classified as either cluster-level, patient-level, or technical deviations. Cluster-level deviations include delayed or unsuccessful implementation of the intervention, inadvertent delivery of the intervention during a control period, prolonged system outages, or incorrect ward assignment. Patient-level deviations include failure to monitor eligible patients or monitoring of patients who do not meet the predefined eligibility criteria. Technical deviations include device malfunctions, communication failures, or command-center outages that affect delivery of the intervention. All randomized patients will remain in the intention-to-treat (ITT) analysis irrespective of protocol deviations. The frequency and nature of deviations will be summarized by randomized treatment group, and major deviations with the potential to affect intervention delivery or outcome ascertainment will be described in the CONSORT flow diagram and explored in prespecified per-protocol sensitivity analyses, if needed.

#### Changes from the protocol

Any difference between this SAP and the final protocol will be tabulated before database lock and approved by the Steering Committee.

**Table S1:** Glossary of terms used in the current report, based on the CONSORT extension for cluster-randomized trials (4)

| Term | Definitions(4) | Applicability to i-PROTECT trial |
|---|---|---|
| Cluster | The unit of randomization. | Hospital ward |
| Control condition | The comparator treatment. | Masked |
| Duration of period | Time (e.g., months) between each step. | 5 periods, each is 2 months |
| Intervention condition | The treatment under evaluation. | |
| Sequence | A sequence of codes defining the order of implementation of the treatment conditions for each cluster. More than one cluster can be allocated to each sequence. | 4 sequences, with 7 clusters in each sequence |
| Step | A planned point at which a cluster or group of clusters crosses from control to intervention. | The period between two steps is 2 months |

**Table S2:** Baseline data.

| | Intention-to-treat cohort | |
|---|---|---|
| | **Intervention (XX)** | **Control (XX)** |
| Age (yr), Median (Q1, Q3) | xx (xx, xx) | xx (xx, xx) |
| Female sex – n (%) | xx (xx) | xx (xx) |
| Admission source – n (%) | | |
| Emergency Room | xx (xx) | xx (xx) |
| Operating Room | xx (xx) | xx (xx) |
| Clinic | xx (xx) | xx (xx) |
| Intensive care unit | xx (xx) | xx (xx) |
| Others | xx (xx) | xx (xx) |
| Admitting ward – n (%) | | |
| Medical | xx (xx) | xx (xx) |
| Surgical | xx (xx) | xx (xx) |
| Oncology | xx (xx) | xx (xx) |
| Mixed (any combination) | xx (xx) | xx (xx) |
| Comorbidities – n (%) | | |
| End-Stage Renal Disease (ESRD) | xx (xx) | xx (xx) |
| Chronic liver disease | xx (xx) | xx (xx) |
| Cancer without metastasis | xx (xx) | xx (xx) |
| Immune-compromised | xx (xx) | xx (xx) |
| Diabetes non-complicated | xx (xx) | xx (xx) |
| Diabetes, complicated | xx (xx) | xx (xx) |
| Congestive heart failure | xx (xx) | xx (xx) |
| Acquired immunodeficiency syndrome | xx (xx) | xx (xx) |
| Moderate to severe chronic kidney disease (CKD) | xx (xx) | xx (xx) |
| Myocardial infarction | xx (xx) | xx (xx) |
| Chronic pulmonary disease | xx (xx) | xx (xx) |
| Peripheral vascular disease | xx (xx) | xx (xx) |
| Stroke or transient ischemic attack | xx (xx) | xx (xx) |
| Dementia | xx (xx) | xx (xx) |
| Hemiplegia or paraplegia | xx (xx) | xx (xx) |
| Connective tissue disease | xx (xx) | xx (xx) |
| Peptic ulcer disease | xx (xx) | xx (xx) |
| Mild liver disease | xx (xx) | xx (xx) |
| Moderate to severe liver disease | xx (xx) | xx (xx) |
| Charlson Comorbidity Index | xx (xx) | xx (xx) |
| Patients post-ICU discharge | | |
| Patients with CCRT activation | | |
| Patients with NEWS-2 of 5 and above | | |

***** no documented infection or infection source is defined as no ICD-10AU for pneumonia, urinary tract infection, skin and soft tissue infection, intra-abdominal infection or other infections.

**Table S3:** Intervention data.

| | Intervention (XX) | Control (XX) |
|---|---|---|
| Patients post-ICU discharge | xx (xx) | xx (xx) |
| % placed on i-PROTECT | xx (xx) | xx (xx) |
| Patients with CCRT activation | xx (xx) | xx (xx) |
| % placed on i-PROTECT | xx (xx) | xx (xx) |
| Patients with NEWS-2 of 5 and above | xx (xx) | xx (xx) |
| % placed on i-PROTECT | xx (xx) | xx (xx) |
| Total placed on i-PROTECT | | |
| Patients with alerts – n (%) | xx (xx) | xx (xx) |
| Alert count per patient- Median (Q1, Q3) | xx (xx, xx) | xx (xx, xx) |
| Time to first alert - Median (Q1, Q3) | xx (xx, xx) | xx (xx, xx) |
| Type of alert | | |
| Tachypnea | xx (xx) | xx (xx) |
| Bradypnea | xx (xx) | xx (xx) |
| Tachycardia | xx (xx) | xx (xx) |
| Bradycardia | xx (xx) | xx (xx) |
| Desaturation | xx (xx) | xx (xx) |
| Alert acknowledgment by nurse | | |
| Percent acknowledged – n (%) | xx (xx) | xx (xx) |
| Time to acknowledgment | xx (xx, xx) | xx (xx, xx) |
| 0-15 minutes – n (%) | xx (xx) | xx (xx) |
| 16-60 minutes – n (%) | xx (xx) | xx (xx) |
| >60 minutes – n (%) | xx (xx) | xx (xx) |
| Action | | |
| Physician informed | xx (xx) | xx (xx) |
| No action needed | xx (xx) | xx (xx) |
| False alert | xx (xx) | xx (xx) |
| Other | xx (xx) | xx (xx) |
| Duration of monitoring | xx (xx, xx) | xx (xx, xx) |

**Table S4:** Outcomes.

| | Intervention (XX) | Control (XX) | RR (95% CI) | p-value | FDR |
|---|---|---|---|---|---|
| Composite endpoint of 30-day in-hospital mortality, cardiac arrest, requirement of vasopressor or intubation -n (%) | xx (xx) | xx (xx) | xx.x (xx.x, xx.x) | 0.xx | 0.xx |
| **Secondary Categorical Outcomes** | | | | | |
| All-cause in-hospital mortality within 30 days– n (%) | xx (xx) | xx (xx) | xx.x (xx.x, xx.x) | 0.xx | 0.xx |
| Cardiac arrest within 30 days – n (%) | xx (xx) | xx (xx) | xx.x (xx.x, xx.x) | 0.xx | 0.xx |
| Requirement of vasopressor within 30 days– n (%) | xx (xx) | xx (xx) | xx.x (xx.x, xx.x) | 0.xx | 0.xx |
| Requirement of intubation within 30 days – n (%) | xx (xx) | xx (xx) | xx.x (xx.x, xx.x) | 0.xx | 0.xx |
| Transfer to ICU within 30 days – n (%) | xx (xx) | xx (xx) | xx.x (xx.x, xx.x) | 0.xx | 0.xx |
| Critical Care Response team (CCRT) activation within 30 days – n (%) | xx (xx) | xx (xx) | xx.x (xx.x, xx.x) | 0.xx | 0.xx |
| **Secondary Continuous outcomes** | | | **Beta estimates (95% CI)** | | |
| Hospital length of stay (LOS), censored at 90 days (days), median (Q1, Q3) | xx (xx, xx) | xx (xx, xx) | xx.x (xx.x, xx.x) | 0.xx | 0.xx |
| ICU-free days in the first 30 days , median (Q1, Q3) | xx (xx, xx) | xx (xx, xx) | xx.x (xx.x, xx.x) | 0.xx | 0.xx |

CI: confidence interval, FDR: false discovery rate, ICU: intensive care unit, Q1: first quartile, Q3: third quartile, RR: relative risk

**Table S5:** Subgroup Analysis.

| | Intervention (XX) | Control (XX) | RR (95% CI) | p-value | Interaction p | FDR |
|---|---|---|---|---|---|---|
| Age=< 65 years | xx (xx) | xx (xx) | xx.x (xx.x, xx.x) | 0.xx | 0.xx | 0.xx |
| Age >65 years | xx (xx) | xx (xx) | xx.x (xx.x, xx.x) | 0.xx | | |
| Admitted to a medical ward | xx (xx) | xx (xx) | xx.x (xx.x, xx.x) | 0.xx | 0.xx | 0.xx |
| Admitted to a surgical ward | xx (xx) | xx (xx) | xx.x (xx.x, xx.x) | 0.xx | 0.xx | 0.xx |
| Admitted to an oncology ward | xx (xx) | xx (xx) | xx.x (xx.x, xx.x) | 0.xx | 0.xx | 0.xx |
| Admitted to a mixed ward | | | | | | |
| Chronic cardiopulmonary disease | xx (xx) | xx (xx) | xx.x (xx.x, xx.x) | 0.xx | 0.xx | 0.xx |
| No chronic cardiopulmonary disease | xx (xx) | xx (xx) | xx.x (xx.x, xx.x) | 0.xx | | |

CI: confidence interval, RR: relative risk, FDR: false discovery rate
*documented infection source includes ICD-10AU for pneumonia, urinary tract infection, skin and soft tissue infection, intra-abdominal infection or other infections.

**References**

1. Hemming K, Taljaard M, McKenzie JE, Hooper R, Copas A, Thompson JA, Dixon-Woods M, Aldcroft A, Doussau A, Grayling M *et al*: **Reporting of stepped wedge cluster randomised trials: extension of the CONSORT 2010 statement with explanation and elaboration**. *BMJ* 2018, **363**:k1614.

### REFERENCES

1. International Council for Harmonisation of Technical Requirements for Pharmaceuticals for Human Use (ICH). E9 Statistical Principles for Clinical Trials. London: ICH; 1998. Available at: https://www.ich.org/page/efficacy-guidelines.
2. Moher D, Hopewell S, Schulz KF, Montori V, Gotzsche PC, Devereaux PJ, et al. CONSORT 2010 explanation and elaboration: updated guidelines for reporting parallel group randomised trials. BMJ. 2010;340:c869.
3. Schulz KF, Altman DG, Moher D, Group C. CONSORT 2010 statement: updated guidelines for reporting parallel group randomised trials. BMJ. 2010;340:c332.
4. Hemming K, Taljaard M, McKenzie JE, Hooper R, Copas A, Thompson JA, et al. Reporting of stepped wedge cluster randomised trials: extension of the CONSORT 2010 statement with explanation and elaboration. BMJ. 2018;363:k1614.
5. Hussey MA, Hughes JP. Design and analysis of stepped wedge cluster randomized trials. Contemp Clin Trials. 2007;28(2):182-91.
6. Shakeshaft A, Doran C, Petrie D, Breen C, Havard A, Abudeen A, et al. The effectiveness of community action in reducing risky alcohol consumption and harm: a cluster randomised controlled trial. PLoS Med. 2014;11(3):e1001617.
7. . McGowan L, Goodman M, https://wwwlexjansencom/nesug/nesug13/118_Final_Paperpdf Accessed on Aug 15-2021.
8. Thompson JA, Hemming K, Forbes A, Fielding K, Hayes R. Comparison of small-sample standard-error corrections for generalised estimating equations in stepped wedge cluster randomised trials with a binary outcome: A simulation study. Stat Methods Med Res. 2021;30(2):425-39.
9. Kenward MG, Roger JH. Small sample inference for fixed effects from restricted maximum likelihood. Biometrics. 1997;53(3):983-97.
10. . LITTLE, RJA , and D B RUBIN (1987) Statistical Analysis with Missing Data New York: John Wiley & Sons.
11. Thijs H, Molenberghs G, Michiels B, Verbeke G, Curran D. Strategies to fit pattern-mixture models. Biostatistics. 2002;3(2):245-65.
12. Benjamini Y, Hochberg Y. Controlling the false discovery rate: a practical and powerful approach to multiple testing. Journal of the Royal statistical society: series B (Methodological). 1995;57(1):289-300.
